# What outcomes are important when evaluating people with hand scars? Results of an international clinician and researcher survey

**DOI:** 10.1101/2023.04.25.23289079

**Authors:** Donna L. Kennedy, Tracy Chism-Balangue, Lucy Dereham, Dominic Furniss

**Author notes:** **Corresponding author:** Dr Donna L. Kennedy, Therapy Department, Charing Cross Hospital, Fulham Palace Road London, W6 8RF United Kingdom.

## Abstract

**Background:** Outcome evaluation in people with hand and wrist scars is not standardised. To improve clinical care and research rigour, the authors are developing a core outcome set (COS) for the evaluation of hand scars. This survey identified what international clinicians and academics consider important outcomes for inclusion when evaluating people with hand scarring.

**Methods:** An electronic survey was developed, peer reviewed and disseminated via professional networks and social media. Professionals of diverse clinical backgrounds and geographical location with experience in the evaluation of hand and wrist scar outcomes in adults were invited to participate. The survey opened in February and closed in May 2022.

**Results:** 162 participants, representing all World Health Organisation geographical regions, completed the survey. 32% of respondents reported using standardised scar patient reported outcome measures (PROMs); 31% using standardised scar clinician reported outcome measures (CROMs). In assessing physical symptoms of scar, sensitivity/hypersensitivity received the highest importance rating, and 36 additional physical symptom constructs were added as free text items by 72 participants. Regarding physical characteristic of scar, evaluation of adhesions was rated as most important and 19 additional physical characteristics were reported by 21 participants. Active range of motion was rated the most important impairment evaluation. In the domain of psychological impact of scarring, satisfaction with scarring and appearance acceptability were rated equally important. Sixty percent of participants reported using a standardised patient-reported outcome measure (PROM) for evaluating the functional impact of scars in the hand and wrist.

**Discussion:** This survey identified items for inclusion in the first round of a stakeholder Delphi consensus study, to agree a COS for the evaluation of hand and wrist scars. Frequency of importance ratings for evaluation constructs were determined to gauge the priorities of survey participants, not to exclude constructs. The disparate outcomes reported by free text within outcome domains highlights the lack of an agreed scar evaluation taxonomy, an important consideration for future consensus work. 107 (66%) of survey respondents consented to contact regarding further scar outcome evaluation consensus work, highlighting this work as a clinical priority.

## BACKGROUND

Our skin is an adaptive barrier protecting us from a harsh environment. When skin is wounded by injury or surgery, scarring results. While all scars may be burdensome, hand scars may be especially deleterious, resulting in altered physical appearance, causing unpleasant or painful symptoms, interfering with activities of daily living and social participation and negatively impacting mental health (1). The significant burden posed by hand scaring was recently highlighted by a British Society for Surgery of the Hand - James Lind Alliance Priority Setting Partnership, where treatment to improve scar and fibrosis formation following hand surgery or trauma was identified as a top ten research priority (2).

Recommendations suggest a comprehensive scar evaluation should include physical characteristics, cosmetic appearance, and symptoms including the impact of scar on activity, social participation, and quality of life (3). While numerous scar evaluation tools exist, at present there is no universally accepted clinician-completed or patient-reported outcome measure (PROM) (4).

Importantly, a recent review of scar assessment methods concluded that current tools fail to capture the impact of scarring on patients’ function, psychosocial health, and quality of life (5). The impact of scarring is multidimensional; therefore, one measure may not adequately capture the outcome domains of importance to people with scars. However, at present, a core outcome set (COS), or agreed standardized set of outcomes, for the evaluation of hand and wrist scars is lacking (6).

To address this gap, we undertook the development of a stakeholder consensus derived hand and wrist scar core outcome measurement set (7). A hand and wrist scar COS will promote standardisation, support evidence synthesis and underpin improvements in clinical research quality, transparency, and rigour (8). The COS development process includes a review of the literature (9) a survey of international clinicians and researchers (reported here), focus groups with people with experience of hand scarring and will conclude with a stakeholder Delphi study (10).

Because scar evaluation is not standardised, we reviewed current practice in evaluation and reporting of scar outcomes in hand and wrist clinical research from inception to December 2022 (9). This enabled the identification of scar associated domains of interest, heterogeneity in outcome measurement and discordance in taxonomy. It was identified that six different standardised scar outcome measures were reported by 25% of studies however only 7% of studies utilised a patient-reported measure. Scar symptoms were the most frequently reported outcome domain; but taxonomy was inconsistent, outcomes lacked working definitions required for generalisability and outcome measurement was disparate and underreported. Nineteen different measures of scar appearance and structure were reported by 51% of studies however only 23% of those were patient-reported. Person-centred domains including scar acceptability, mental health impact, and social participation were rarely reported.

Reviewing the state-of-the-art in the evaluation and reporting of hand scar outcomes in clinical research enabled us to identify *what happens in practice*. In contrast, the aim of this survey was to identify what clinicians and researchers report *should happen* relative to the evaluation of hand scar outcomes. It was anticipated this process, in conjunction with the results of patient and public focus groups, would ensure saturation of scar associated domains, constructs, or outcomes, to inform the first round of our subsequent Delphi consensus study.

### Research question

What scar outcome domains are considered important by clinicians and researchers with experience of hand and wrist scar evaluation in adults?

## METHODS

### Survey design and piloting

An electronic survey was developed using Qualtrics, an online survey tool. Content included scar associated outcomes and domains identified in data synthesis (literature review and qualitative evidence(1)). Survey content was curated to determine which scar associated outcome domains were considered important by clinicians and researchers with experience of evaluating people with hand and wrist scars. Participants were additionally able to report items in free text. The survey was anonymised; however, participants were asked to provide an email address if they wished to receive a copy of the survey results or consented to contact regarding future stakeholder activities. Relevant demographic data was collected including profession, geographical location, self-reported level of expertise in scar evaluation, clinical populations treated or assessed and whether the survey participant was a clinician, academic or clinical academic. The survey was submitted to the British Association of Hand Therapists (BAHT) clinical evidence committee (CEC) for pilot testing and peer review, prior to dissemination.

Participants were asked to rate the importance of evaluating identified measures within specific scar outcome domains, including the use of patient or clinician reported standardised scar outcome measures, scar physical symptoms, physical characteristics, impairment measures, hand function and psychological impact. For each domain and at the conclusion of the survey, participants were invited to free text additional responses. Domain queries were worded, for example, “Do you think it is important to evaluate any of the following physical characteristics of scar?”. Response options and relevant numeric values were definitely not (1); probably not (2); neutral/unsure (3); probably yes (4) and definitely yes (5). For transparency, survey content is available online (11). Data was analysed and reported as the frequency of participants reporting each response category for each item.

Professionals of all clinical backgrounds and geographical location of practice were invited to participate. The survey was disseminated using Facebook, LinkedIn and Twitter social media platforms and by professional organisations including BAHT, the British Society for Surgery of the Hand (BSSH) Research Committee, the British Association of Plastic, Aesthetic and Reconstructive Surgeons (BAPRAS) and the Reconstructive Surgical Trials Network (RSTN). Snowball recruitment was utilised, whereby participants were requested to share the survey with colleagues and professional networks. The survey opened on February 11th 2022 and closed on May 21st 2022.

## RESULTS

### Participant demographics

One hundred sixty-two survey responses were included in data analysis. Participants identified their professional group as Hand Therapist (35%); Occupational Therapist (22%); Hand Surgeon (Orthopaedics) (18%); Physiotherapist (10%); Hand Surgeon (Plastics)(9%); Nurse (1%); Orthotist/Prosthetist (1%) and other (< 1%) (Burns & Trauma Surgeon; Orthopaedics; Patient and Public Representative to the British Society for Surgery of the Hand; Physical Medicine and Rehabilitation (PM&R); Physiatrist; Massage Therapist; Burn Surgeon; Hand Surgeon (General Surgery)). Survey respondents participated from all World Health Organisation (WHO) geographical regions (**Fig 1**), with 48% of participants from the United Kingdom and 27% from the United States.

**Fig 1.**
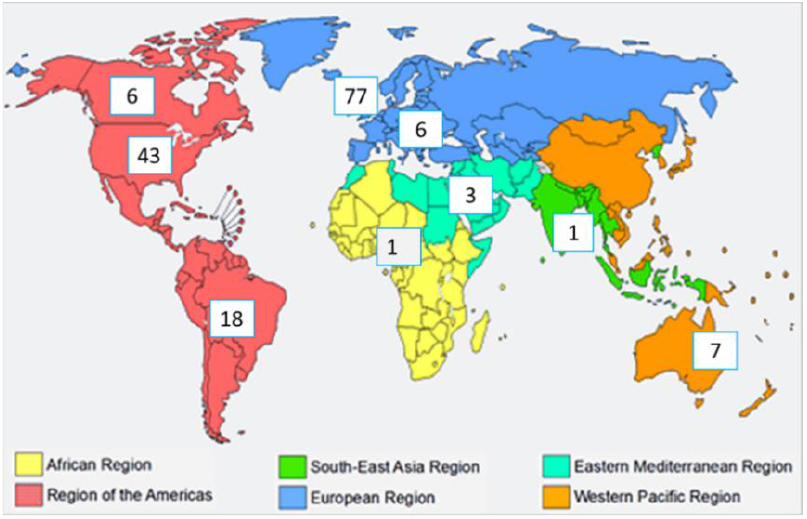
Location of survey participants by World Health Organisation (WHO) geographical regions (https://www.who.int/)

One hundred and seven (66%) survey respondents consented to contact regarding further scar outcome evaluation consensus work.

Participant self-rated level of experience / expertise in scar evaluation was novice (7%); intermediate (50%) and expert (41%). Professional role was reported as clinician (74%); clinical academic (22%) and academic / researcher (4%). Most participants reported experience of evaluating scars in patients with various clinical conditions (69%) (**Table 1**).

**Table 1.** Participant reported clinical cohort of evaluation expertise

| Clinical population | % |
| --- | --- |
| Burns | 9 |
| Elective (planned) hand surgery (i.e., carpal tunnel surgery) | 11 |
| Orthopaedic trauma (i.e., fracture open reduction & internal fixation) | 5 |
| Trauma (i.e. flexor tendon repair) | 4 |
| Complex trauma (i.e., traumatic, open multi-structure injuries) | 3 |
| Combination of the above | 69 |

### Survey Data

#### PROMs

In response to the query, ‘do you use any of the following standardised patient-reported outcome measures (PROMs) for the evaluation of hand and wrist scars?’, 68% of participants reported they do not use standardised scar PROMs. Fifteen percent of participants reported using the Patient and Observer Scar Assessment Scale (POSAS)(12); 3% the Scar-Q (13); 2% the Patient Scar Assessment Questionnaire (PSAQ) (14) ; and 1% the Patient-Reported Impact of Scars Measure (PRISM) (15). Free text responses included CARe Burn Scales (16) (1%) and the Brisbane Burn Scar Impact Profile (17) (1%). Nine percent of participants added free text items which were not scar PROMs, for example, the modified Kapanji test (18).

#### CROMs

Regarding the use of standardised scar clinician-reported outcome measures (CROMs) for the evaluation of hand and wrist scars, 69% of participants reported not using these tools. Twenty-four percent of respondents reported using the Vancouver Scar Scale (19), and 1% each the Manchester Scar Scale (20), Matching Assessment of Scars and Photographs (21), and Stony Brook Scar Evaluation Scale (22). Additional free text responses included the Observer Scale of the POSAS (12) (3%); Visual Analogue Appearance Score (VAAS) (n=1); Derriford Appearance Score (23) (n=1); and the Japan Scar Scale 2015 (24) (n=1).

#### Physical Symptoms

The importance of assessing the physical symptoms of scar, including hypersensitivity, pain, hyperesthesia, allodynia, dysesthesia and itch were rated by participants (**Fig 2**). Sensitivity / hypersensitivity received the highest importance rating with 98% of participants reporting hypersensitivity should definitely or probably be evaluated. This was closely followed by pain, with 96% reporting pain should definitely or probably be evaluated. Of note, for all included physical symptoms, more than 75% of respondents reported each construct should probably or definitely be included in scar evaluation. Additional free text scar physical symptoms outcomes of importance are reported in **Supplementary Data 1**.

**Fig 2.**
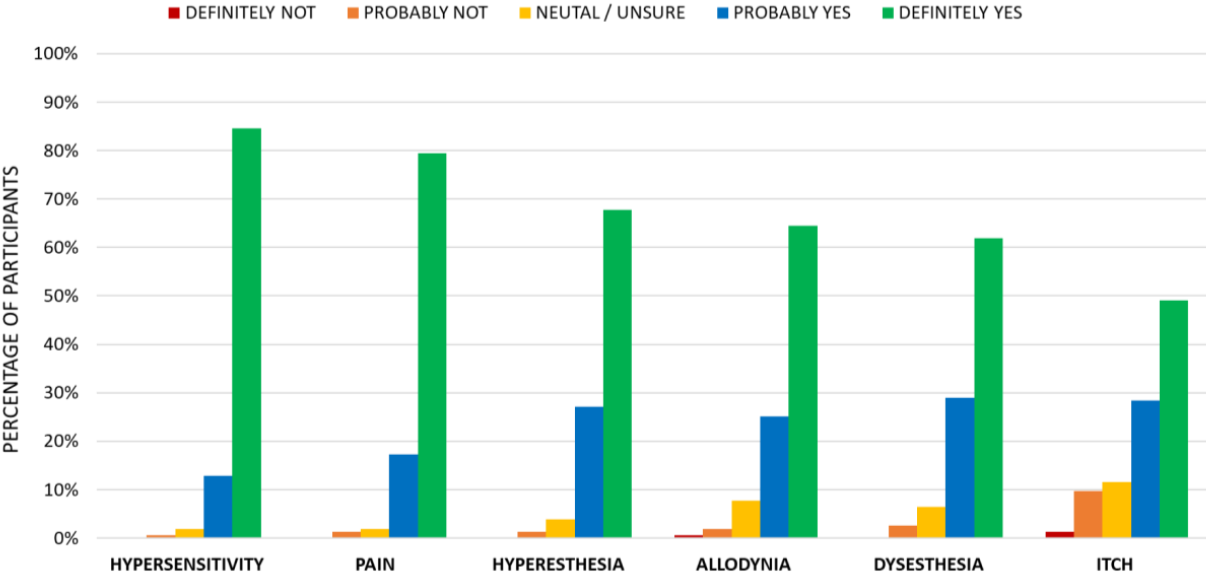
Participant-rated importance of evaluating physical symptoms of scars.

#### Physical characteristics

Evaluating adhesions, as a physical characteristic of scar, received the highest rating importance rating with 97% of respondents stating that adhesions should definitely or probably be evaluated.

This was followed by pliability (93% definitely or probably include); colour (88% definitely or probably include) and thickness (88% definitely or probably include) **(Fig 3)**. In contrast, less than 50% of respondents reported that dryness, relief, shine or sweating should definitely be included in scar outcome evaluation. Additional free text scar physical characteristics for inclusion in evaluation are reported in **Supplementary Data 2**.

**Fig 3.**
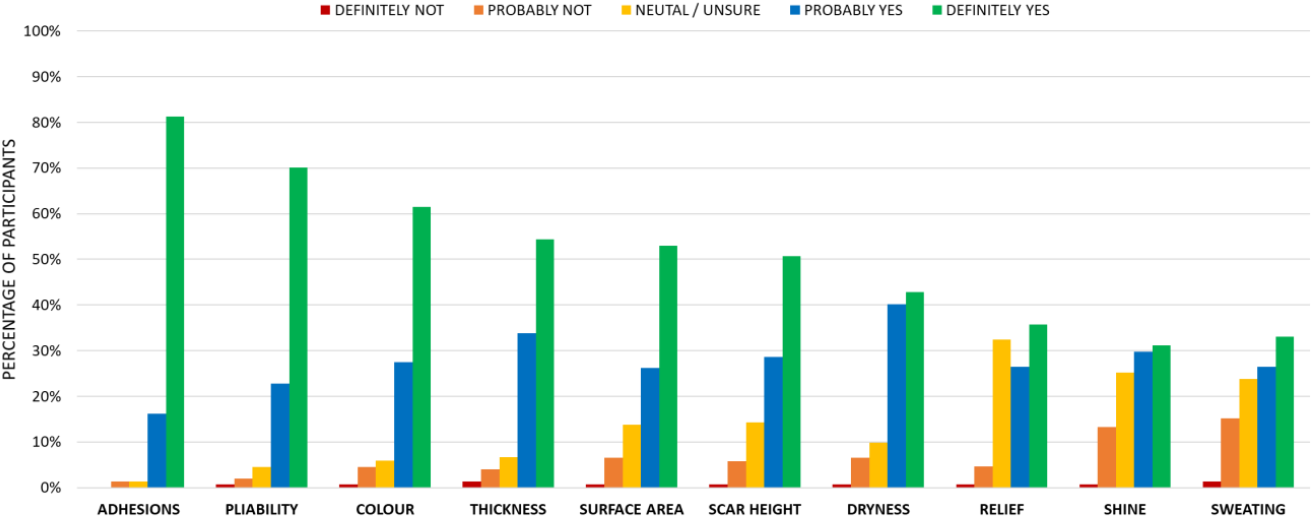
Participant rated importance of evaluating physical characteristics of scars.

#### Impairment Measures

Participants rated the importance of assessing impairment measures when evaluating hand scars, including range of motion, strength, sensation and dexterity. Evaluating active range of motion (AROM) was rated as the most important outcome construct, with 98% of respondents stating that AROM should definitely or probably be evaluated. This was followed closely by passive range of motion in terms of relative importance (97% of respondents rated definitely or probably include). **(Fig 4)**. In contrast, less than 40% of respondents thought it was definitely important to evaluate grip strength, sensory detection threshold, pinch strength, manual muscle testing or innervation density. Additional free text impairment measures, alphabetically, included dynamometry (n=1); functional impairment (n=2); grip (n=1); light touch (n=1); pinch (n=1); sharp (n=1); verbal pain scale (n=1) and vibration (n=1).

**Fig 4.**
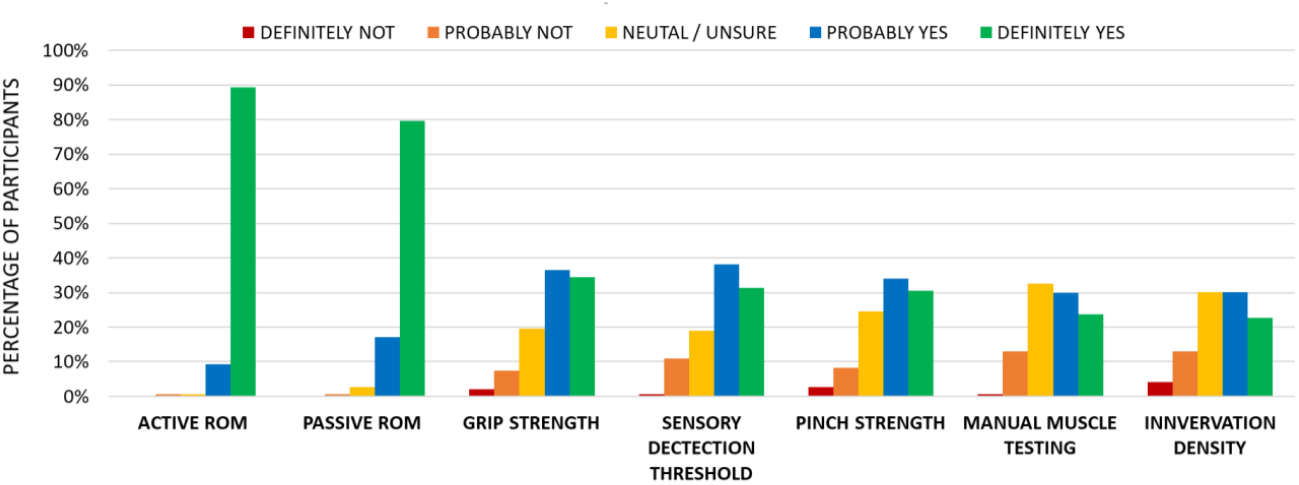
Participant-rated importance of evaluating impairment measures during hand scars evaluation.

#### Mental Health Impact

Survey participants rated the importance of evaluating various psychological and mental health impacts of scarring (**Fig 5**). Satisfaction with scarring and appearance acceptability were equally rated as the most important outcome constructs, with 95% of respondents stating that scar satisfaction or appearance acceptability should definitely or probably be evaluated. Additionally, 93% of participants noted that behaviour compensation should definitely or probably be evaluated. In contrast, less than 50% of participants thought that self-esteem (49%), anxiety (47%), PTSD (45%), stigmatisation (46%), self-confidence (46%) or anger (35%) should definitely be evaluated. Additional free text constructs included close family’s response to scar (n=1) and rumination, focus on the scar (n=1).

**Fig 5.**
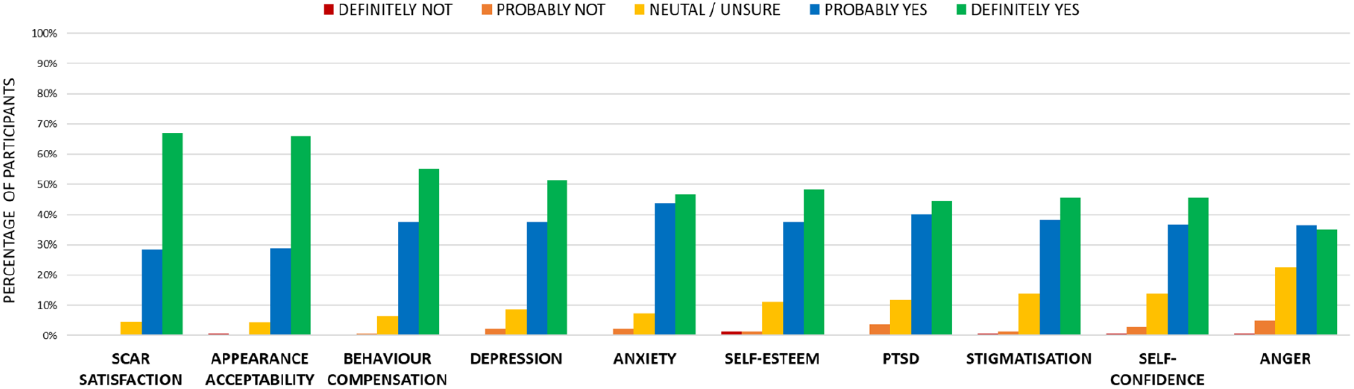
Participant-rated importance of evaluating psychological or mental health impact of hand scars.

#### Hand Function PROMs

Ninety-seven (60%) of participants reported using a standardised patient-reported outcome measure (PROM) when evaluating the functional impact of scars in the hand and wrist. Of those PROMs used by respondents, the Disabilities of the Arm, Shoulder and Hand Outcome Measure (DASH) (25) was used most often (n=64; 66%), followed by the Patient Rated Wrist and Hand Evaluation (26) (n=34; 35%), Michigan Hand Outcome Measure (27) (n=10; 10%), Patient Specific Functional Scale (28) (n=10; 10%), Boston Carpal Tunnel Questionnaire (29) (n=9; 9%), Canadian Occupational Performance Measure (30) (n=7; 7%), Patient Evaluation Measure (31) (n=5; 5%) and the QuickDash (32) (n=5; 5%). Additional free text PROM hand function measures included the iHaND (33) (n=1) and the Upper Extremity Functional Index (34) (n=1).

#### Other Measures

Participants were invited, at the survey conclusion, to write as free text any additional outcome domains or constructs not previously identified that they felt should be included in the evaluation of hand and wrist scars. Free text items included ability of client to massage scar independently (n=1); balance/symmetry (n=1); capillary refill time (n=1); CARe burn scales (n=1); composite range of motion (n=1); duration of scar (n=1); functional alterations (n=1); functional limitations (n=1); oedema (n=1); pain at rest and movement and touch evoked (n=1); participation (n=1); patient expectations (n=1); photographic tracking (n=1); psychosocial impact (n=1); quality of life measures (EQ5D-5L or SF36/VF36) (n=1); return to pre-injury function (n=2); sensory alterations (n=1) and timing of evaluation (n=1).

## DISCUSSION

The overarching aim of this work was to identify items for inclusion in the first round of a stakeholder Delphi consensus study, to agree a core outcome set (COS) for the evaluation of people with hand and wrist scars. This survey specifically aimed to identify what experienced international clinicians and academics deem to be important outcomes for inclusion in the evaluation of people with hand scarring.

Clinicians and academics representing all of the World Health Organisation geographical regions participated. Sixty-eight percent of respondents reported not using standardised scar patient reported outcome measures (PROMs), whilst 69% reported not using standardised scar clinician reported outcome measures (CROMs). In assessing the physical symptoms of scar, sensitivity/hypersensitivity received the highest importance rating, and 36 additional physical symptom constructs were added as free text items by 72 participants. Regarding physical characteristic of scar, the evaluation of adhesions was rated as most important; active range of motion was rated as the most important impairment evaluation. In the domain of mental health or psychological impact of scarring, satisfaction with scarring and appearance acceptability were equally important outcomes. Sixty percent of participants reported using a standardised patient-reported outcome measure (PROM) for evaluating the functional impact of scars in the hand and wrist, most commonly the Disabilities of the Arm, Shoulder and Hand Outcome Measure (DASH).

Frequency ratings were determined to gauge the priorities of survey participants for evaluation constructs, not to exclude constructs with lower ratings. Survey and free text items will inform the first round Delphi survey. The disparate outcomes reported by free text within outcome domains highlights the lack of an agreed scar evaluation taxonomy, an important consideration for future consensus work. One hundred and seven (66%) survey respondents consented to contact regarding further scar outcome evaluation consensus work, highlighting this as a clinical priority.

## Data Availability

All data produced in the present work are contained in the manuscript.

## Declarations

### 1. Conflicting interests

The authors report no conflicts

### 2. Funding

DLK was supported by the NIHR Imperial Biomedical Research Centre (BRC) and an Imperial Clinical Academic Training Office (CATO) Postdoctoral Bridging Fellowship.

### 3. Informed consent

Survey participation was taken as informed consent

### 4. Ethical approval

Not applicable

### 5. Contributorship

DLK, TCB and DF researched literature, conceived and disseminated the survey. DLK and DF drafted and revised the survey. DLK and LD performed data analysis. All authors contributed to drafting the manuscript and approved the final version of the manuscript.

## 6. Acknowledgements

We would like to thank our survey participants who generously gave of their time and shared their expertise

## 7. Disclosures

This report is independent research and the views expressed in this publication are those of the authors and not necessarily those of the NHS, the National Institute for Health Research, or the Department of Health.

## Supplementary Data

**Supplementary Data 1.** Survey respondents additional free text scar physical symptoms of importance (alphabetically)

| Physical symptom | n = |
| --- | --- |
| active range of motion | 2 |
| adhesions/adherence | 7 |
| aesthetic outcome/appearance | 9 |
| bulk | 1 |
| burning | 2 |
| colour | 6 |
| contracture | 4 |
| cosmesis | 1 |
| depth | 1 |
| dysfunction | 1 |
| elasticity/flexibility | 2 |
| firmness | 1 |
| functional impact | 3 |
| glide/mobility/tethering | 2 |
| height | 2 |
| hypertrophy | 1 |
| keloid | 1 |
| length | 1 |
| measurement | 1 |
| movement limiting effect | 1 |
| neuropathic pain | 1 |
| pigmentation | 1 |
| pliability | 1 |
| psychological element | 1 |
| satisfaction | 1 |
| sensory effect on work & ADL | 1 |
| shape | 2 |
| size/dimensions | 3 |
| soft tissue fibrosis | 1 |
| tethering | 1 |
| texture | 1 |
| thickness | 3 |
| tightness /stretching | 3 |
| vascularity | 2 |
| width | 1 |
| n = number of participants free texting cited measure |  |

**Supplementary Data 2.** Survey respondents additional free text scar physical characteristics of importance (alphabetically)

| **Physical characteristic** | **n=** |
| --- | --- |
| abnormalities | 1 |
| blend with surrounding area | 1 |
| contractures | 2 |
| fragile epidermal layer | 1 |
| hardness | 1 |
| impact on motion – tension visually | 1 |
| induration | 1 |
| location | 1 |
| nodules | 1 |
| passive range of motion | 1 |
| pigmentation | 2 |
| presence of stitches | 1 |
| raised | 1 |
| refill | 1 |
| retraction | 1 |
| smoothness | 1 |
| surrounding skin condition | 1 |
| tightness of scar with adjacent joints movement | 1 |
| vascularity | 1 |
| n = number of participants free texting cited measure |  |

## Notes

### Competing Interest Statement

The authors have declared no competing interest.

### Funding Statement

Donna L. Kennedy was supported by the NIHR Imperial Biomedical Research Centre (BRC) and an Imperial Clinical Academic Training Office (CATO) Postdoctoral Bridging Fellowship.

